# A cross-sectional study of the association between COVID-19 infection and psychological distress in Japanese workers

**DOI:** 10.1101/2023.04.08.23288312

**Authors:** Hirofumi Tesen, Yusuke Konno, Seiichiro Tateishi, Kosuke Mafune, Mayumi Tsuji, Akira Ogami, Tomohisa Nagata, Ryutaro Matsugaki, Reiji Yoshimura, Yoshihisa Fujino, the CORoNaWork Project

## Abstract

The COVID-19 pandemic infected many people worldwide with SARS-CoV2. Psychological distress is one of the sequelae reported to occur in many of those infected (Choutka et al., 2022). We investigated the association between personal experience of COVID-19 infection and psychological distress in Japan. A total of 18,560 persons participated in the original survey, conducted in December 2020. After excluding unreliable responses, data from 14,901 persons who participated in a follow-up survey in December 2022-were included in the analysis. Odds ratios (ORs) were estimated by univariate and multiple logistic regression analysis with history of COVID-19 infection as the independent variable and presence of psychological distress as the dependent variable.

This results showed that the experience of COVID-19 infection is associated with psychological distress. Moreover, most cases of mental distress among those who experienced COVID-19 infection can be at least partly explained by a perception of unfair treatment.

## 1. Introduction

COVID-19, which was identified in 2019 and spread worldwide, has caused enormous numbers of infections worldwide in repeated waves of transmission. In Japan, as of February 2023 a total of more than 33 million people have been infected (Ministry of health, Labor and Welfare of Japan, 2023). The global fatality rate reportedly decreased from 8.5% February 2020 to 0.27% August 2022, attributable to greater vaccine uptake, effectiveness of treatment, acquisition of immunity, and progressive weakening of the virus (Horita et al., 2023). The breakdown of acute severity of 457 patients in Japan discharged after treatment for COVID-19 was 84.4% mild, 12.7% moderate, and 2.9% severe. Symptoms in the acute phase included olfactory impairment (7.7%), fatigue (6.6%), dyspnea (3.9%), impaired taste (3.5%), and cough (2.4%). Fewer than 5% of patients reported still having symptoms after one year, but in some cases the latter can become chronic (Tsuzuki et al., 2022).

The persistence of symptoms in some patients is referred to as “post-COVID-19 conditions” or “long COVID”. The World Health Organization (WHO) defined post-COVID-19 conditions as symptoms lasting at least 2 months and not explained by other diseases. Symptoms that appear after the acute phase include memory impairment, poor concentration, and depression. These symptoms remained in 10% of patients after 6 month and in about 5% after 1 year (Miyazato et al., 2022). Recently, reports on long COVID have been increasing. An analysis of U.S. medical cost reimbursement data showed that 27,698 (32%) of 87,337 patients with COVID-19 had sequelae, with increasing the likelihood of long COVID including respiratory failure, hypertension, memory impairment, renal impairment, and psychiatric disorders (Ettman et al., 2022). In addition, a study of cognitive dysfunction in elderly COVID-19 patients reported an increased risk of developing cognitive dysfunction six months later compared to non-COVID-19 patients with other severe or mild illness (Liu et al., 2022).

Long COVID may lead to various psychiatric and neurological symptoms. Studies have reported increased number of patients presenting with depressive and anxiety symptoms after COVID-19 infection, with depressive symptoms particularly more likely to persist (Fancourt et al. 2022). In a report from Japan, fatigue (13%), poor concentration (8%), sleep disturbance (7%), and memory impairment (7%) were reported as residual symptoms 1 year after COVID-19 diagnosis (Nakagawara et al., 2021). In a large survey of U.S. veterans, one year after COVID-19 survivors were 46% more likely than controls to have been diagnosed with a psychiatric disorder: the latter included depression, suicidal ideation, anxiety, sleep disorders, and cognitive dysfunction (Xie et al., 2022). Analyses of data from 236,379 cases, mainly in the U.S., revealed that 34% of COVID-19 patients developed a psychiatric disorder within six months, and that after recovery, psychiatric disorders such as depression and anxiety were more common than influenza and other respiratory infections (Taquet et al., 2021).

Psychological manifestations of long COVID-19 have been related to not only biological mechanisms associated with the infection, but also to psychological and socioeconomic factors. In a review of the psychological effects of isolation at home or in quarantine facilities during the COVID-19 pandemic, most studies were shown to report psychological effects such as trauma, confusion, and anger (Brooks et al., 2020). It is widely known that deprivation of social interaction due to isolation has negative psychological effects (Valtorta et al., 2012). In Japan, early COVID-19 infection cases were subjected to 2-week quarantine reduced to 10 days from December 2020.

Although the number of cases of infection in Japan continues to rise, there are no reports on psychological problems after the experience of infection. We hypothesized that mental problems after infection experience would be related to experience of discrimination. Indeed, discrimination, inhibition, and social exclusion occur in the context of various infectious diseases, with COVID-19 infected individuals and their close contacts reportedly more vulnerable to unemployment and economic disadvantages (Wilson et al., 2020). Such negative consequences are thought to be linked to post-infection psychiatric symptoms. We therefore examined whether discriminatory experiences affect post-infection psychological problems among COVID-19-infected individuals.

## 2. Methods

### 2.1. Study Design and Participants

The original study consisted of a cross-sectional cohort survey of 27,036 participants, conducted in December 2020. The present study focused on the 18,560 who participated in the December 2022 follow-up survey.

The Collaborative Online Research on the Novel-Coronavirus and Work (CoroNaWork) Project is an Internet-based study investigating the health status of workers during the COVID-19 pandemic in Japan in December 2020. Details of the protocol for this survey are available elsewhere; the basic design is summarized below (Fujino et al. 2021).

For the baseline survey in December 2020 approximately 600,000 pre-registered individuals were invited to participate, and approximately 55,000 met the participation criteria. They had to be employed at the time of the survey and between the ages of 20-60. Gender, work type (office work or not), and region of residence were taken into account to ensure uniformity among participants. Five regions of residence were defined, based on COVID-19 infection rates for the 47 prefectures in Japan. In all, 33,302 people completed in the survey, with data from 27,036 retained for analysis after excluding those who submitted untrustworthy responses (e.g., completion times extremely short (less than 6 minutes), reporting height below 140 cm, reporting weight below 30 kg, or giving inconsistent answers to multiple identical questions).

A total of 18,560 participants were included in both the baseline survey (conducted between December 22 and 25, 2020 and the follow-up conducted between December 1 and 5, 2022). Individuals with a Kessler-6 score of 10 or higher at baseline were excluded (N=3,269). We also excluded 291 individuals who were not employed at follow-up. Finally, 14,901 persons were included in the analysis. The study was conducted with the approval of the Ethics Committee of the University of Occupational and Environmental Health, Japan (Approval No. R2-079). Informed consent was obtained through the website.

### 2.2. Measurements at baseline

Participants were asked to answer Yes or No to the following questions regarding their experience with COVID-19 infection after the baseline survey (all questions were in Japanese):

1. Has been diagnosed as infected with a SARS-CoV2
2. Has tested positive for SARS-CoV2 on a PCR test
3. Has tested positive for SARS-CoV2 antigens

Respondents answering “Yes ”to any of the three questions were categorized as having experienced Covid-19 infection.

Other factors investigated in the questionnaire included age, sex, marital status, job type, educational level, finances, alcohol consumption, and number of employees at the workplace; these were considered as confounding factors. We also asked the question: “Do you think you have been treated unfairly?”

### 2.3. Assessment of psychological distress

The Kessler 6-item psychological distress scale (K6, Japanese version) was used to measure psychological distress (Kessler et al., 2002). The following six questions were asked:

“During the past 30 days, how often have you felt (1. irritable, 2. hopeless, 3. restless, 4. unwell, 5. fractured, 6. unworthy of yourself)?”

Each item was rated on a scale of 0 to 4 (0. never, 1. a little, 2. sometimes, 3.usually, 4. always) according to frequency.

The K6 Japanese version’s under the curve domain for mood or anxiety disorders in DSM-IV was 0.94, with solid validity (Furukawa et al., 2008). Cutoffs vary, with some investigators regarding scores of 5 to 12 points as moderate and 13 or more as severe psychological distress (Sakurai et al., 2011). The Japanese Ministry of Health, Labour and Welfare’s “National Survey of Living Conditions ”treats a score of 10 or higher as psychological distress equivalent to a mood or anxiety disorder; we also used a score of 10 or higher as the cutoff.

### 2.4. Statistical analysis

Univariate and multiple logistic regression analyses were used to estimate odds ratios (ORs) with COVID-19 infection experience as the independent variable and presence or absence of psychological distress as the dependent variable. Given the difficulty in differentiating infection related effects and potential stress vulnerability, we focused on participants with a baseline K6 score of below 10.

In the multivariate model, we adjusted for age and sex (model 1). We further adjusted for marital status, finances, educational level, alcohol consumption, job type, number of employees at the workplace, and baseline K6 score (model 2). We further adjusted for the response to the question “Do you feel you have been treated unfairly? ”as a variable (model 3).

P-values of less than 0.05 were considered statistically significant. We used Stata (Stata Statistical Software: Release 16. College Station, TX: StataCorp LLC.) for all analyses.

## 3. Results

Table 1 shows the general characteristics of the 14,901 people who provided data for analysis. Of these, 166 (1.1%) had a history of infection. Age, marital status, occupation, education, and alcohol consumption items were similar between those with and without a history of infection. In contrast, univariate analysis showed that those with a history of infection were more likely to be male, had higher K6 scores, and more likely to feel they had suffered economically or had been treated unfairly.

**Table 1.** The characteristics of participants who have experienced infection.

| Characteristics | non-infection<br>n=14735 (%) | infection<br>n=166(%) |
| --- | --- | --- |
| Age (years), mean (SD) | 49.2 (9.7) | 47.7 (11.6) |
| Sex, male | 8649 (58.7%) | 112 (67.5%) |
| Marriage status |  |  |
| married | 8741 (59.3%) | 103(62.0%) |
| Job type |  |  |
| Mainly desk work | 7712 (52.3%) | 81 (48.8%) |
| Mainly work involving communicating with people | 3556 (24.1%) | 50 (30.1%) |
| Mainly labor | 3467 (23.5%) | 35 (21.1%) |
| Educational background |  |  |
| Junior high school | 189 (1.3%) | 1 (0.6%) |
| High school | 3780 (25.7%) | 47 (28.3%) |
| University, graduate school, vocational school, junior college | 10766 (73.1%) | 118 (71.1%) |
| Number of employees in the workplace |  |  |
| <10 | 3596 (24.4%) | 37 (22.3%) |
| <100 | 3745 (25.4%) | 44 (26.5%) |
| <1000 | 3631 (24.6%) | 44 (26.5%) |
| >1000 | 3763 (25.5%) | 41 (24.7%) |
| Finance |  |  |
| very difficult | 1428 (9.7%) | 25 (15.1%) |
| difficult | 3883 (26.4%) | 58 (34.9%) |
| normal | 7092 (48.1%) | 65 (39.2%) |
| comfortable | 1963 (13.3%) | 18 (10.8%) |
| very comfortable | 369 (2.5%) | 0 (0.0%) |
| Current smoke | 4067 (27.6%) | 49 (29.5%) |
| Alcohol consumption |  |  |
| 2 or more days a week | 6491 (44.1%) | 71 (42.8%) |
| Do you think you have been treated unfairly? | 120 (0.8%) | 43 (25.9%) |
| K6 |  |  |
| median (Interquartile Range) | 1.0 (0.0, 4.0) | 3.0 (0.0, 6.0) |
| >5 | 3578 (24.3%) | 58 (34.9%) |

Table 2 shows the Odds Ratio (OR) between infection experience and K6, as estimated by the logistic model. The age-sex adjusted OR for the association between infection experience and higher K6 was significant (OR=2.75, 95%CI 1.85-4.00). The association result was also significant in multivariate analysis model 2 (adjusted for baseline K6) with OR=2.15, 95%CI 1.42-3.26. In model 3 (adjusted for “feeling treated unfairly”), the association between infection experience and a high K6 score was significantly attenuated (OR=1.61, 95%CI 1.02-2.55).

**Table 2.**
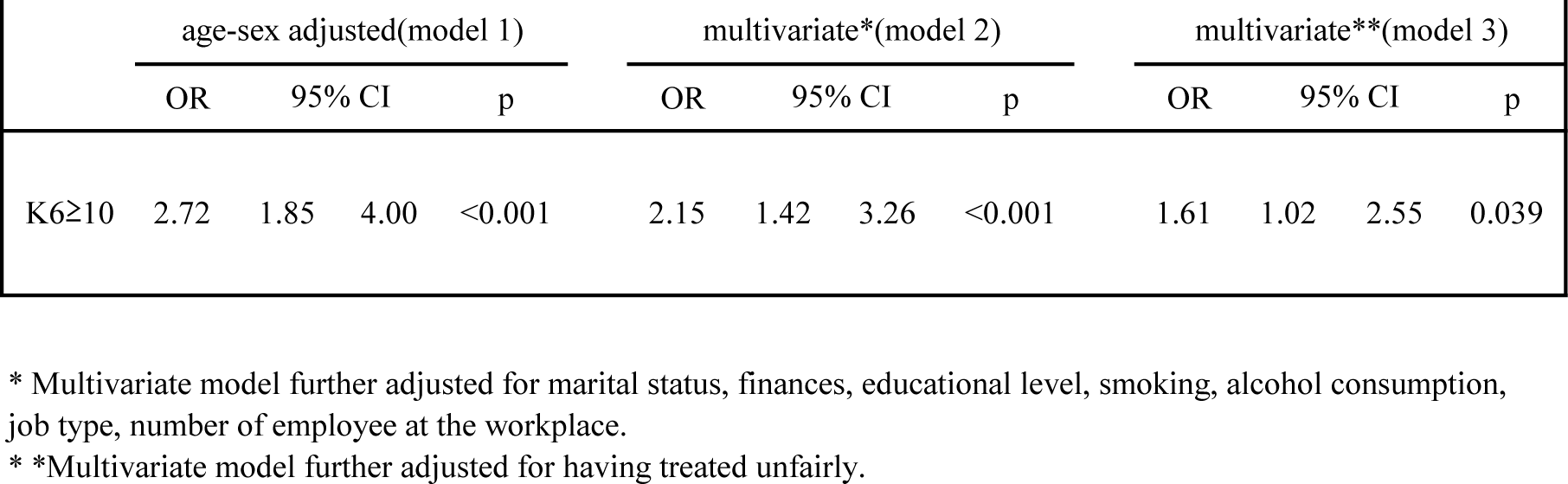
The Association between experience of infection and K6

## 4. Discussion

The follow-up survey revealed that 1.1% of the 14,901 people whose data were analyzed had a history of infection, and that experiencing COVID-19 infection was associated with psychological distress. Moreover, the psychological distress of infected individuals was at least partially explainable by their experience of unfair treatment. To our knowledge, this is the first large-scale study in Japan to investigate the history of COVID-19 infection and psychological distress.

In this study, individuals without psychological distress at baseline were at higher risk of developing psychological distress after experiencing COVID-19 infection. Mental health effects after infection are a typical symptom of long COVID (Fancourt et al., 2022). Neurological and psychiatric symptoms are associated with numerous types of viral infection, but long COVID differs from other viral infections in that symptoms may be unremarkable at the time of infection but develop after a mild or moderate bout (Choutka et al., 2022). Long COVID is thought to be caused not only by biological mechanisms associated with infection, but also by socio-psychological factors (Wadman et al., 2022; Brooks et al., 2020; Valtorta et al., 2012).

Concerning biological mechanisms, symptoms of long COVID, including reduced ability to speak, remember, or concentrate have been reported after infection. These cognitive sequelae are referred to as “brain fog,” and recently proposed to be related to brain inflammation in depression (Beurel et al., 2020; Steardo et al., 2020). Activation of antibodies and deposition of amyloid aggregates are among the possible underlying mechanisms (Apple et al., 2022; Fernandez et al., 2022; Charnley et al., 2022). It has also been noted that the symptoms of long COVID are similar to those experienced by patients undergoing cancer chemotherapy and those with myalgic encephalomyelitis, chronic fatigue syndrome, and mast cell activation syndrome. It has been suggested that mast cells stimulated by pathogenic viruses or stress stimuli may activate microglia and induce hypothalamic inflammation (Theoharis et al., 2021).

Concerning socio-psychological factors, in Japan, people with COVID-19 were originally required by law to be isolated for about 2 weeks in a hospital, convalescent hotel, or at home. Restrictions on home life and employment due to isolation are known to have psychological consequences (Valtorta et al., 2012). A study of Chinese youths reported increased suicidal ideation and self-harm after the strict COVID-19 social isolation policy was enforced (Zhang et al., 2020). Moreover, the recommendation of social distancing as a countermeasure against the spread of COVID-19 led to spread of other problems worldwide, such as loneliness and isolation. Whereas spending time with family members during a pandemic reduces loneliness (Fujii, et al.2022), social distancing among family members may exacerbate feeling of loneliness. In addition, the increasing number of infected persons in Japan resulted in, scattered cases of infection within families. Fear that family members may become infected is associated with psychological distress (Wang et al., 2021). During a pandemic the family serves as a buffer, while the endangerment of family well-being can alter resilience (Prime et al., 2020).

There may be socioeconomic disadvantages for infected individuals, as those who have been infected or had close contact with the disease are reported to be at higher risk of unemployment (Nagata et al., 2021). Reduced income during the pandemic, and insecurity surrounding employment and finances are reported to increase the risk of depression, anxiety, and insomnia, requiring early psychological support and some form of intervention (Li et al., 2020; Wilson et al., 2020). Furthermore, during the COVID-19 pandemic, delayed return to work was found to negatively impact mental health (Shi et al., 2020).

It is against this general background that experiences of discrimination and stigma appear relevant. Under a pandemic, stigma is not merely possible, but a common phenomenon (Xiang et al., 2020), and stigma associated with COVID-19 has already been identified as an important stressor (Hamouche et al., 2020; Giorgi et al., 2020). It is directed at infected persons and those who come in contact with them (Bai et al., 2004; Brooks et al., 2020). Essential workers may be required to continue working even during a pandemic despite strains on mental health due to constantly being on guard against unexpected contact and resulting stigma and discrimination (Sasaki et al., 2021). For example, in Japan, a social problem arose early in the COVID-19 epidemic when children of nurses and other health care workers were denied access to daycare centers. Complaints about discrimination related to COVID-19 are common in the workplace (Choi et al., 2021; Kantamneni et al., 2020). Because they may increase feeling of shame and guilt in the target individual, discrimination and stigma can lead to reduced feelings of self-worth (Corrigan et al., 2002). These findings are supported by the results of the present study: the association between experience of infection and K6 score was greatly attenuated when adjusted for discriminatory experiences.

The present study suggests that secondary feelings of being treated unfairly, or so-called stigma issues, have a significant role in the emergence of psychological distress. Reports on COVID-19 infection and risk of psychiatric disorders continue to grow in number, and in parallel with the search for biological mechanisms, clarification of psychosocial aspects is required. COVID-19-infected individuals may experience a double burden of clinical distress as well as stigmatizing social disadvantage; they experience decreased self-worth and disconnection from society following COVID-19 infection, and secondary environmental changes that follow infection. Even after symptoms are in remission, medium-to long-term care, including social support, is needed. To address stigma and preserve the patient’s self-respect, the patient should not be socially isolated, should enjoy positive relationships with family and friends, and have appropriate interactions with others (Lubkin et al., 2007). In addition, WHO has expressed concern about infodemics with respect to COVID-19 (Novel Coronavirus.situation report, 2020). It is important to provide correct information in real time to prevent stigma exacerbation through misinformation (Hamouche et al., 2022). Measures to adequately address stigmatizations may help to counter post-infection psychological distress.

### 4.1. Limitation

The following limitations of this study should be taken into consideration: First, information on infection experience was obtained through self-reports. Clinical characteristics and actual severity of infection were not ascertained. However, several questions related to infection experience were administered to check for internal validity. Second, psychological distress was determined only by participants’ K6 ratings, so it was not possible to evaluate the severity of mental illness as accurately as with other measurement tools, such as the Depression Rating Scale. Third, precise information about the timing of the infection experience was not requested, so it is possible that people who had been infected for less than two months were included.

Fourth, given the a cross-sectional nature of the study, the temporal relationship between infection experience and psychological distress issues is unknown. However, the sample was from the population that participated in the cohort study, and as we excluded respondents with high K6 scores at baseline, we know that all participants had normal K6 scores prior to infection.

### 4.2. Conclusion

In this study, the experience of infection during the COVID-19 pandemic was found to be associated with psychological distress. This association may be influenced by secondary changes such as discrimination and stigma following infection.

## Credit statement

YF: The chairperson of the study group, Conceptualization, Methodology, Formal analysis, Editing; HT: Original draft, Writing and Editing.

## Data availability

Requests for the original data presented in the study can be directed to the corresponding author.

## Data Availability

All data produced in the present study are available upon reasonable request to the corresponding author

## Acknowledgements

We thank other members of the CORoNaWork Project.

## References

1. Apple, A.C., Oddi, A., Peluso, M.J., Asken, B.M., Henrich, T.J., Kelly, J.D., Pleasure, S.J., Deeks, S.G., Allen, I.E., Martin, J.N., Ndhlovu, L.C., Miller, B.L., Stephens, M.L., Hellmuth, J., 2022. Risk factors and abnormal cerebrospinal fluid associate with cognitive symptoms after mild COVID-19. Ann. Clin. Transl. Neurol. 9, 221–226.

2. Bai, Y., Lin, C.-C., Lin, C.-Y., Chen, J.-Y., Chue, C.-M., Chou, P., 2004. Survey of stress reactions among health care workers involved with the SARS outbreak. Psychiatr. Serv. 55, 1055–1057.

3. Beurel, E., Toups, M., Nemeroff, C.B., 2020. The bidirectional relationship of depression and inflammation: double trouble. Neuron 107, 234–256.

4. Brooks, S.K., Webster, R.K., Smith, L.E., Woodland, L., Wessely, S., Greenberg, N., Rubin, G.J., 2020. The psychological impact of quarantine and how to reduce it: rapid review of the evidence. Lancet 395, 912–920.

5. Charnley, M., Islam, S., Bindra, G.K., Engwirda, J., Ratcliffe, J., Zhou, J., Mezzenga, R., Hulett, M.D., Han, K., Berryman, J.T., Reynolds, N.P., 2022. Neurotoxic amyloidogenic peptides in the proteome of SARS-COV2: potential implications for neurological symptoms in COVID-19. Nat. Commun. 13, 1–11.

6. Choi, C., Kulkarni, M.P., n.d. In one month, STOP AAPI HATE receives almost 1500 incident reports of verbal harassment, shunning, and physical assaults. Asian Pacific Policy & …. Press Releases.

7. Choutka, J., Jansari, V., Hornig, M., Iwasaki, A., 2022. Unexplained post-acute infection syndromes. Nat. Med. 28, 911–923.

8. Corrigan, P.W., Watson, A.C., 2002. Understanding the impact of stigma on people with mental illness. World Psychiatry 1, 16–20.

9. Ettman, C.K., Cohen, G.H., Abdalla, S.M., Sampson, L., Trinquart, L., Castrucci, B.C., Bork, R.H., Clark, M.A., Wilson, I., Vivier, P.M., Galea, S., 2022. Persistent depressive symptoms during COVID-19: a national, population-representative, longitudinal study of U.S. adults. The Lancet Regional Health - Americas 5, 100091.

10. Fancourt, D., Steptoe, A., Bu, F., 2022. Psychological consequences of long COVID: comparing trajectories of depressive and anxiety symptoms before and after contracting SARS-CoV-2 between matched long- and short-COVID groups. Br. J. Psychiatry 1–8.

11. Fernández-Castañeda, A., Lu, P., Geraghty, A.C., Song, E., Lee, M.-H., Wood, J., O’Dea, M.R., Dutton, S., Shamardani, K., Nwangwu, K., Mancusi, R., Yalçın, B., Taylor, K.R., Acosta-Alvarez, L., Malacon, K., Keough, M.B., Ni, L., Woo, P.J., Contreras-Esquivel, D., Toland, A.M.S., Gehlhausen, J.R., Klein, J., Takahashi, T., Silva, J., Israelow, B., Lucas, C., Mao, T., Peña-Hernández, M.A., Tabachnikova, A., Homer, R.J., Tabacof, L., Tosto-Mancuso, J., Breyman, E., Kontorovich, A., McCarthy, D., Quezado, M., Vogel, H., Hefti, M.M., Perl, D.P., Liddelow, S., Folkerth, R., Putrino, D., Nath, A., Iwasaki, A., Monje, M., 2022. Mild respiratory COVID can cause multi-lineage neural cell and myelin dysregulation. Cell 185, 2452–2468.e16.

12. Fujii, R., Konno, Y., Tateishi, S., Hino, A., Tsuji, M., Ikegami, K., Nagata, M., Yoshimura, R., Matsuda, S., Fujino, Y., 2021. Association between time spent with family and loneliness among Japanese workers during the COVID-19 pandemic: A cross-sectional study. Front. Psychiatry 12, 786400.

13. Fujino, Y., Ishimaru, T., Eguchi, H., Tsuji, M., Tateishi, S., Ogami, A., Mori, K., Matsuda, S., 2021. Protocol for a nationwide internet-based health survey of workers during the COVID-19 pandemic in 2020. J. UOEH 43, 217–225.

14. Furukawa, T.A., Kawakami, N., Saitoh, M., Ono, Y., Nakane, Y., Nakamura, Y., Tachimori, H., Iwata, N., Uda, H., Nakane, H., Watanabe, M., Naganuma, Y., Hata, Y., Kobayashi, M., Miyake, Y., Takeshima, T., Kikkawa, T., 2008. The performance of the Japanese version of the K6 and K10 in the World Mental Health Survey Japan. Int. J. Methods Psychiatr. Res. 17, 152–158.

15. Giorgi, G., Lecca, L.I., Alessio, F., Finstad, G.L., Bondanini, G., Lulli, L.G., Arcangeli, G., Mucci, N., 2020. COVID-19-related mental health effects in the workplace: a narrative review. Int. J. Environ. Res. Public Health 17. https://doi.org/10.3390/ijerph17217857

16. Hamouche, S., 2020. COVID-19 and employees’ mental health: stressors, moderators and agenda for organizational actions. Emerald Open Research 2. https://doi.org/10.35241/emeraldopenres.13550.1

17. Horita, N., Fukumoto, T., 2023. Global case fatality rate from COVID-19 has decreased by 96.8% during 2.5 years of the pandemic. J. Med. Virol.95, https://doi.org/10.1002/jmv.28231

18. Kantamneni, N., 2020. The impact of the COVID-19 pandemic on marginalized populations in the United States: A research agenda. J. Vocat. Behav. 119, 103439.

19. Kessler, R.C., Andrews, G., Colpe, L.J., Hiripi, E., Mroczek, D.K., Normand, S.L.T., Walters, E.E., Zaslavsky, A.M., 2002. Short screening scales to monitor population prevalences and trends in non-specific psychological distress. Psychol. Med. 32, 959–976.

20. Liu, Y.-H., Chen, Y., Wang, Q.-H., Wang, L.-R., Jiang, L., Yang, Y., Chen, X., Li, Y., Cen, Y., Xu, C., Zhu, J., Li, W., Wang, Y.-R., Zhang, L.-L., Liu, J., Xu, Z.-Q., Wang, Y.-J., 2022. One-year trajectory of cognitive changes in older survivors of COVID-19 in Wuhan, China: A longitudinal cohort study. JAMA Neurol. 79, 509–517.

21. Li, X., Lu, P., Hu, L., Huang, T., Lu, L., 2020. Factors associated with mental health results among workers with income losses exposed to COVID-19 in China. Int. J. Environ. Res. Public Health 17. https://doi.org/10.3390/ijerph17155627

22. Lubkin, I.M., Larsen, P.D., 2006. Chronic illness: Impact and interventions. Jones & Bartlett Learning. Ministry of health, Labor and Welfare of Japan, 2023. Retrieved at 15 February 2023 from, https://covid19.mhlw.go.jp/extensions/public/en/index.html

23. Miyazato, Y., Tsuzuki, S., Morioka, S., Terada, M., Kutsuna, S., Saito, S., Shimanishi, Y., Takahashi, K., Sanada, M., Akashi, M., Kuge, C., Osanai, Y., Tanaka, K., Suzuki, M., Hayakawa, K., Ohmagari, N., 2022. Factors associated with development and persistence of post-COVID conditions: A cross-sectional study. J. Infect. Chemother. 28, 1242–1248.

24. Nakagawara, K., Namkoong, H., Terai, H., Masaki, K., Tanosaki, T., Shimamoto, K., Lee, H., Tanaka, H., Okamori, S., Kabata, H., Chubachi, S., Ikemura, S., Kamata, H., Yasuda, H., Kawada, I., Ishii, M., Ishibashi, Y., Harada, S., Fujita, T., Ito, D., Bun, S., Tabuchi, H., Kanzaki, S., Shimizu, E., Fukuda, K., Yamagami, J., Kobayashi, K., Hirano, T., Inoue, T., Kagyo, J., Shiomi, T., Ohgino, K., Sayama, K., Otsuka, K., Miyao, N., Odani, T., Oyamada, Y., Masuzawa, K., Nakayama, S., Suzuki, Y., Baba, R., Nakachi, I., Kuwahara, N., Ishiguro, T., Mashimo, S., Minematsu, N., Ueda, S., Manabe, T., Funatsu, Y., Koh, H., Yoshiyama, T., Saito, F., Ishioka, K., Takahashi, S., Nakamura, M., Goto, A., Harada, N., Kusaka, Y., Nakano, Y., Nishio, K., Tateno, H., Edahiro, R., Takeda, Y., Kumanogoh, A., Kodama, N., Okamoto, M., Umeda, A., Hagimura, K., Sato, T., Miyazaki, N., Takemura, R., Sato, Y., Takebayashi, T., Nakahara, J., Mimura, M., Ogawa, K., Shimmura, S., Negishi, K., Tsubota, K., Amagai, M., Goto, R., Ibuka, Y., Hasegawa, N., Kitagawa, Y., Kanai, T., Fukunaga, K., 2021. Comprehensive and long-term surveys of COVID-19 sequelae in Japan, an ambidirectional multicentre cohort study: study protocol. BMJ Open Respir. Res. 8, e001015.

25. Novel Coronavirus.situation report, 2020. Retrieved at 15 June 2020 from, https://www.who.int/docs/default-source/coronaviruse/situation-reports/20200202-sitrep-13-ncov-v3.pdf

26. Prime, H., Wade, M., Browne, D.T., 2020. Risk and resilience in family well-being during the COVID-19 pandemic. Am. Psychol. 75, 631–643.

27. Sakurai, K., Nishi, A., Kondo, K., Yanagida, K., Kawakami, N., 2011. Screening performance of K6/K10 and other screening instruments for mood and anxiety disorders in Japan. Psychiatry Clin. Neurosci. 65, 434–441.

28. Sasaki, N., Kuroda, R., Tsuno, K., Imamura, K., Kawakami, N., 2021. Deterioration in mental health under repeated COVID-19 outbreaks greatest in the less educated: A cohort study of Japanese employees. J. Epidemiol. 31, 93–96.

29. Shi, L., Lu, Z.-A., Que, J.-Y., Huang, X.-L., Liu, L., Ran, M.-S., Gong, Y.-M., Yuan, K., Yan, W., Sun, Y.-K., Shi, J., Bao, Y.-P., Lu, L., 2020. Prevalence of and risk factors associated with mental health symptoms among the general population in China during the coronavirus disease 2019 pandemic. JAMA Netw Open 3, e2014053.

30. Steardo, L., Jr, Steardo, L., Verkhratsky, A., 2020. Psychiatric face of COVID-19. Transl. Psychiatry 10, 261.

31. Taquet, M., Luciano, S., Geddes, J.R., Harrison, P.J., 2021. Bidirectional associations between COVID-19 and psychiatric disorder: retrospective cohort studies of 62 354 COVID-19 cases in the USA. Lancet Psychiatry 8, 130–140.

32. Theoharides, T.C., Cholevas, C., Polyzoidis, K., Politis, A., 2021. Long-COVID syndrome-associated brain fog and chemofog: Luteolin to the rescue. Biofactors 47, 232–241.

33. Tsuzuki, S., Miyazato, Y., Terada, M., Morioka, S., Ohmagari, N., Beutels, P., 2022. Impact of long-COVID on health-related quality of life in Japanese COVID-19 patients. Health Qual. Life Outcomes 20, 125.

34. Valtorta, N., Hanratty, B., 2012. Loneliness, isolation and the health of older adults: do we need a new research agenda? J. R. Soc. Med. 105, 518–522.

35. Wadman, M., 2022. Lasting impact of infection extends to the brain. Science 375, 707.

36. Wang, Y., Di, Y., Ye, J., Wei, W., 2021. Study on the public psychological states and its related factors during the outbreak of coronavirus disease 2019 (COVID-19) in some regions of China. Psychol. Health Med. 26, 13–22.

37. Wilson, J.M., Lee, J., Fitzgerald, H.N., Oosterhoff, B., Sevi, B., Shook, N.J., 2020. Job insecurity and financial concern during the COVID-19 pandemic are associated with worse mental health. J. Occup. Environ. Med. 62, 686–691.

38. Xiang, Y.-T., Yang, Y., Li, W., Zhang, L., Zhang, Q., Cheung, T., Ng, C.H., 2020. Timely mental health care for the 2019 novel coronavirus outbreak is urgently needed. Lancet Psychiatry 7, 228–229.

39. Xie, Y., Xu, E., Al-Aly, Z., 2022. Risks of mental health outcomes in people with covid-19: cohort study. BMJ 376, e068993.

40. Zhang, L., Zhang, D., Fang, J., Wan, Y., Tao, F., Sun, Y., 2020. Assessment of mental health of Chinese primary school students before and after school closing and opening during the COVID-19 pandemic. JAMA Netw Open 3, e2021482.

